# Survival and neurologic outcomes after re-irradiation in children with diffuse midline glioma and diffuse intrinsic pontine glioma

**DOI:** 10.64898/2026.05.29.26354429

**Authors:** Tina Vaziri, Dhara Vyas, Mohannad Alhumaid, Calixto-Hope Lucas, Melike Guryildirim, Lindsay Kilburn, Robyn D. Gartrell, Michael A. Koldobskiy, Eric Raabe, Kenneth Cohen, Matthew Ladra, Sahaja Acharya

## Abstract

**Background:** Re-irradiation (reRT) is increasingly offered following progression in diffuse intrinsic pontine glioma (DIPG) and diffuse midline glioma (DMG), though optimal patient selection remains a challenge. This study evaluated clinical outcomes after reRT in a contemporary cohort of patients with DIPG/DMG.

**Methods:** Patients ≤26 years old with DMG/DIPG treated with radiation therapy between 2011-2025 were retrospectively reviewed. Primary endpoints included overall survival (OS2) and progression-free survival (PFS2), measured from first progression, and change in neurologic symptoms after reRT. Survival was estimated using Kaplan–Meier methods, with Cox proportional hazards modeling for prognostic factors.

**Results:** Fifty-eight patients were included; 37 (63.8%) underwent reRT. Tumors were predominantly pontine (74.1%). ReRT was associated with improvement in motor function (51.4% vs. 9.5%, p=0.002), cranial nerve function (29.7% vs. 4.8%, p=0.044), and gait ataxia (35.1% vs. 9.5%, p=0.059). Median OS2 and PFS2 were improved with reRT (OS2: 9.67 vs. 2.57 months, *p*<0.001; PFS2: 5.63 vs. 1.57 months, *p*<0.001). OS2 was independently associated with reRT (HR 0.27, *p*<0.0001), pontine location (HR 2.94, *p*=0.004), and steroid use at progression (HR 4.12, *p*=0.001). PFS2 was independently associated with reRT (HR 0.23, *p* <.0001) and distant pattern of failure (HR 2.83, *p*=.037). Among reRT patients, non-pontine location was associated with improved OS2 (*p*=0.02), and local failure was associated with improved PFS2 (*p*=0.003).

**Conclusion:** ReRT was associated with neurologic improvement and prolonged survival. Patients with non-pontine tumors or local-only failure might derive the greatest benefit. Prospective studies are warranted to define optimal dose/fractionation and refine patient selection.

**Key Points:**

- Reirradiation in patients with DMG/DIPG improved overall survival and progression free survival
- Reirradiation improved neurologic outcomes
- Local-only failure and non-pontine tumor location may derive greatest benefit from reirradiation

**Importance of the Study:** In patients with diffuse intrinsic pontine glioma and diffuse midline glioma, progression after initial treatment remains nearly universal, and salvage therapies are limited. Re-irradiation (reRT) is increasingly offered, yet optimal patient selection and neurologic benefit remain unclear. Additionaly, prior studies exploring reRT lack a comparison cohort of patients that did not receive reRT and rarely integrate neurologic or molecular data. This study investigates the impact of reRT in a contemporary cohort of patients with molecular characterization and includes assessment of neurologic impact. We provide evidence that reRT can improve neurologic function and extend survival, particularly in patients with non-pontine tumors or local-only recurrence. By identifying factors associated with better outcomes, our findings help guide individualized decision-making for reRT.

**Lay Summary:** Pediatric patients with aggressive brain tumors, including diffuse intrinsic pontine glioma and diffuse midline glioma, often have tumor recurrence and neurologic decline despite initial treatment. Options when the tumor grows again are limited. This study shows that giving a second course of radiation, called re-irradiation, can help improve neurologic symptoms including movement and coordination, and extend survival. We also identify features like tumor location and site of tumor regrowth that help to predict who may benefit most. These findings can help physicians make informed decisions about using re-irradiation to improve quality of life and outcomes for those facing these challenging tumors.

## Introduction

Diffuse intrinsic pontine glioma (DIPG) is a devastating pediatric brain tumor that primarily arises in the pons and accounts for approximately 80% of brainstem gliomas and 10-15% of all pediatric central nervous system tumors.^1^ DIPG presents with rapid-onset of symptoms including cranial nerve palsies, motor weakness, and cerebellar dysfunction such as ataxia and dysmetria.^2^ Diagnosis is largely clinical and radiographic, with MRI typically demonstrating a non-enhancing, T2-hyperintense lesion diffusely infiltrating the pons.^2^ Biopsies were historically avoided; however, advances in stereotactic techniques have allowed for safer sampling.^3^ Most DIPGs are associated with a histone *H3 p.K27M* mutation and are pathologically classified as Diffuse Midline Glioma (DMG), *H3K27*-altered.^4,5^ These tumors commonly have the hallmark histone *H3* mutation which drives oncogenesis through epigenetic dysregulation. Less common subtypes of DMG can be driven by pathogenic mutation or amplification of *EGFR* or overexpression of *EZHIP*.^4,5^ Although DIPG is anatomically centered in the pons, DMGs can arise in other midline structures such as the thalamus and spinal cord. These non-pontine DMGs often present with location-specific neurologic deficits such as hemiparesis or increased intracranial pressure in thalamic lesions or myelopathy in spinal cord lesions.^6,7^ In contrast to pontine DMGs which predominantly affect pediatric patients, non-pontine DMGs demonstrate a broader age distribution and are more frequently observed in adolescents and young adults.^6,8^ Furthermore, co-occurring mutations in genes such as *TP53*, *ATRX*, and components of the *MAPK* pathway (*NF1, FGFR1, FGFR2, KRAS,* and *BRAF*) may stratify tumor behavior and prognosis.^9^

Despite improvements in diagnostic accuracy and molecular insight, prognosis for DMG/DIPG remains poor due to the tumor’s deep anatomical location and resulting inability to achieve a gross total resection, infiltrative growth, and resistance to current therapies. The median overall survival for pontine DMG is 9-12 months, and fewer than 10% of patients survive beyond two years.^10,11^ Most of the survival data are based on historical reports of DIPG or pontine DMG as data on non-pontine DMG are scarce. Radiation therapy (RT) remains the standard of care for DIPG/DMG, providing temporary symptom relief and a modest extension of survival.^12^ However, all patients inevitably experience tumor progression, highlighting the urgent need for more effective treatments.

To date, randomized clinical trials evaluating systemic therapies for progressive disease have failed to demonstrate a survival benefit.^13,14^ Re-irradiation (reRT) has emerged as a viable option in this context, offering both palliative and survival benefits. A matched-cohort analysis found that reRT extended median survival by 3-6 months.^15^ Meta-analyses and institutional case series have similarly shown palliative benefits, including improved quality of life and motor function.^16,17^ Although not curative, reRT remains one of the few evidence-supported treatment options that can improve quality of life and extend survival in children with progressive DMG/DIPG.

Despite growing interest in reRT for children with DMG and DIPG, existing literature has several limitations. Most published studies include a substantial proportion of patients without biopsy and therefore, largely represent clinically and radiographically defined DIPG rather than molecularly confirmed DMG.^15,16,18,19^ Additionally, it remains unclear whether outcomes following reRT are similar for patients with non-pontine DMG compared with those with DIPG.

To address these gaps, we analyzed a contemporary cohort of patients, including children with DMG arising outside of the pons, the majority of whom have undergone a biopsy. The objectives of this study were to evaluate the association between reRT and overall survival (OS2) and progression-free survival (PFS2) after first progression, and to assess the impact of reRT on neurologic function.

## Materials and Methods

### Study population

The study was approved by the Institutional Review Board at **Anonymized for Review**. We performed a retrospective review of patients aged ≤26 years with a diagnosis of DMG or DIPG treated at **Anonymized for Review** between 2011 and 2025 who completed an initial course of RT and subsequently experienced disease progression. Diagnosis was established radiographically and clinically as DIPG^1^ or by molecular confirmation as DMG *H3K27*-altered. A pathologic diagnosis of DMG required all of the following: (1) a diffuse glioma, (2) loss of *H3K27me3* by immunohistochemistry, (3) midline location and (4) presence of *H3 p.K27M* or *H3 p.K27I*, or presence of a pathogenic mutation or amplification of *EGFR* or overexpression of *EZHIP* or methylation profile of one of the DMG subtypes.

Patients were excluded if they met any of the following criteria: (1) metastatic disease at the time of diagnosis, (2) no documented progression, (3) did not meet diagnostic criteria for DIPG or DMG, (4) did not receive standard RT as first line therapy and (5) patient had less than three months of follow-up after first progression (**Supplemental Figure 1**). Standard RT for first line therapy was defined as 50.4 Gy conventionally fractionated or 39 Gy in 13 fractions as a hypo-fractionated regimen.

### Clinical variables

Data extracted from electronic medical records included patient demographics, date of diagnosis, pathology and molecular reports, neurologic symptoms, RT dose and fractionation, date of progression, imaging findings, and date of death or last follow-up. The date of diagnosis was defined as the date of histopathologic confirmation or, in cases of radiographically diagnosed DIPG, the date of the diagnostic MRI.

### Histopathologic Review and DNA methylation profiling

Immunohistochemistry was performed on formalin-fixed, paraffin-embedded tissue in a Clinical Laboratory Improvement Amendments (CLIA)- certified laboratory using the following antibodies: H3K27me3 (C36B11, cat#9733, Cell Signaling Technology, 1:100) and H3 K27M (EPR18340, cat#ab190631, Abcam, 1:1000). DNA methylation profiling was performed through the National Institute of Health/National Cancer Institute or the Childhood Cancer Data Initiative Molecular Characterization Initiative at Nationwide Children’s Hospital, as previously described.^20,21^ The DKFZ/Heidelberg random forest classification algorithm was used for tumor classification.^22^

### Neurologic Symptom Assessment and Corticosteroid Use

Five neurologic symptoms were abstracted from clinical encounters: motor strength, cranial nerve deficit, dysphagia, gait ataxia, and dysarthria. These were assessed at: (1) initial presentation, (2) first progression, and (3) first follow-up after second line treatment. Each neurologic symptom was defined as present or absent at initial presentation and at first progression. On analysis, each neurologic symptom was treated as a binary variable, and a composite neurologic symptom score (range 0–5) was calculated as the sum of affected domains at each timepoint. Higher scores reflected greater symptomatic burden. Use of corticosteroids was also determined and coded as a binary variable at each visit. At first follow-up after reRT, typically scheduled six to eight weeks after completion of reRT, each neurologic symptom was coded as stable, improved, or worsened based on comparison of symptoms at first progression. For patients who did not receive reRT, neurologic symptoms were assessed 2-3 months after first progression.

### Treatment course and follow-up

The initial treatment was determined by a multidisciplinary team including pediatric neuro-oncologists, neurosurgeons and radiation oncologists, following institutional protocols and patient-specific considerations. All patients received focal RT as part of first line therapy.

Patients were followed clinically as well as radiographically with brain MRI at least every three months following completion of the initial RT course. Disease progression was defined as either radiographic tumor enlargement on MRI,^23^ and/or worsening neurological symptoms attributable to tumor progression, as determined by the clinical team. The decision to offer reRT was individualized, considering factors such as the patient’s performance status, interval since initial RT, and prior response to RT.

### Endpoints

The primary endpoints are OS and PFS. OS1 was defined as the time from the date of initial diagnosis to the date of death or last follow-up. OS2 was defined as the time from the date of first disease progression to the date of death or last follow-up. PFS1 was measured from the date of initial diagnosis to the date of progression, death or last follow-up, whichever occurred first. PFS2 was measured from the date of first disease progression to the date of second progression, death or last follow-up, whichever occurred first.

### Immortal Time Bias

A challenge in evaluating treatments administered shortly after disease progression is that patients must survive long enough to receive the intervention (e.g. reRT) to be included in the treatment group, which can introduce a bias favoring reRT. To mitigate potential immortal time bias, we performed a landmark analysis excluding patients who died within one month of progression, as this interval may not have allowed sufficient time for initiation of reRT.^24^

### Statistical Analysis

Patient and disease characteristics were summarized with descriptive statistics reported as frequencies/percentages or medians with interquartile range (IQR) or range for continuous variables. Baseline characteristics between the reRT and no reRT groups were compared using Fisher’s exact or chi-square tests as appropriate for categorical variables, and continuous variables were compared using the Wilcoxon rank-sum test. The Kaplan-Meier method was used to generate survival curves. Groups were compared using the log-rank test. Prognostic factors associated with OS and PFS were assessed by Cox proportional hazards regression. Variables were included in the multivariate model if the p-value was <0.05 on univariate analysis. A multivariable model was constructed using a stepwise backward selection method. Analysis was performed using Stata v.18 (StataCorp LLC, College Station, TX).

## Results

### Patient, tumor, and treatment characteristics

Fifty-eight patients with a diagnosis of DIPG/DMG who developed disease progression after completing first line RT were included. Among these, 37 patients received reRT at first progression, while 21 patients did not.

Table 1 summarizes the cohort’s clinical and disease characteristics. The median age at diagnosis was 7.7 years (range, 3.2–26.7). Tumors were primarily located in the pons (74.14%), followed by thalamus (22.41%) and midbrain (3.45%). Most patients had pathologic confirmation of DMG (82.75%). Among the 48 patients who had available molecular data, 46 had confirmed *H3 p.K27M* mutations and 2 had a methylation profile consistent with *EZHIP* overexpression subtype of DMG. The median initial RT dose was 54 Gy and majority of patients received conventional fractionation (96.55%). Baseline clinical characteristics were similar between the reRT and no reRT groups although patients diagnosed between 2018-2025 were more likely to undergo reRT (81.08% vs. 52.38%, p =.035) (**Table 1**).

**Table 1.**
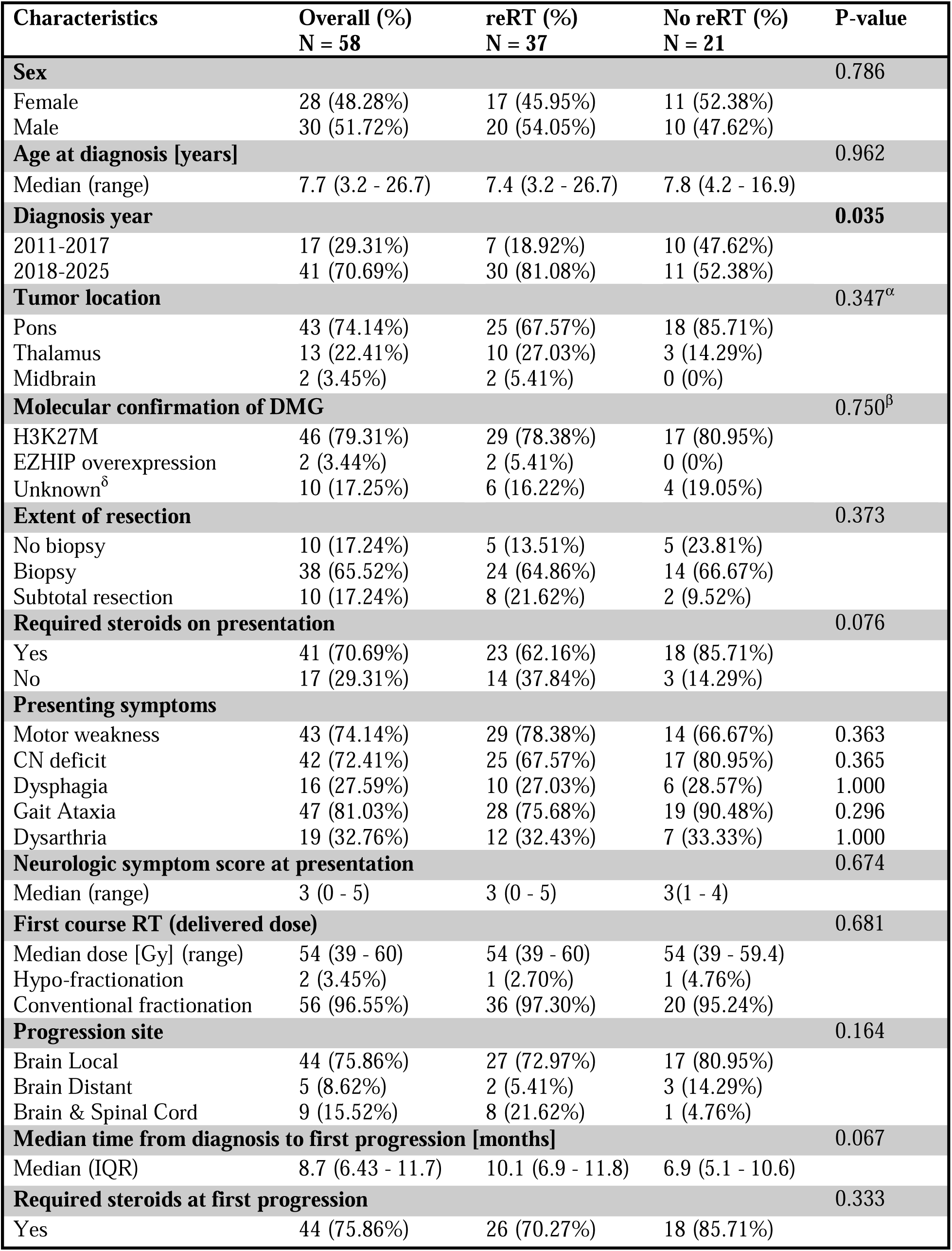
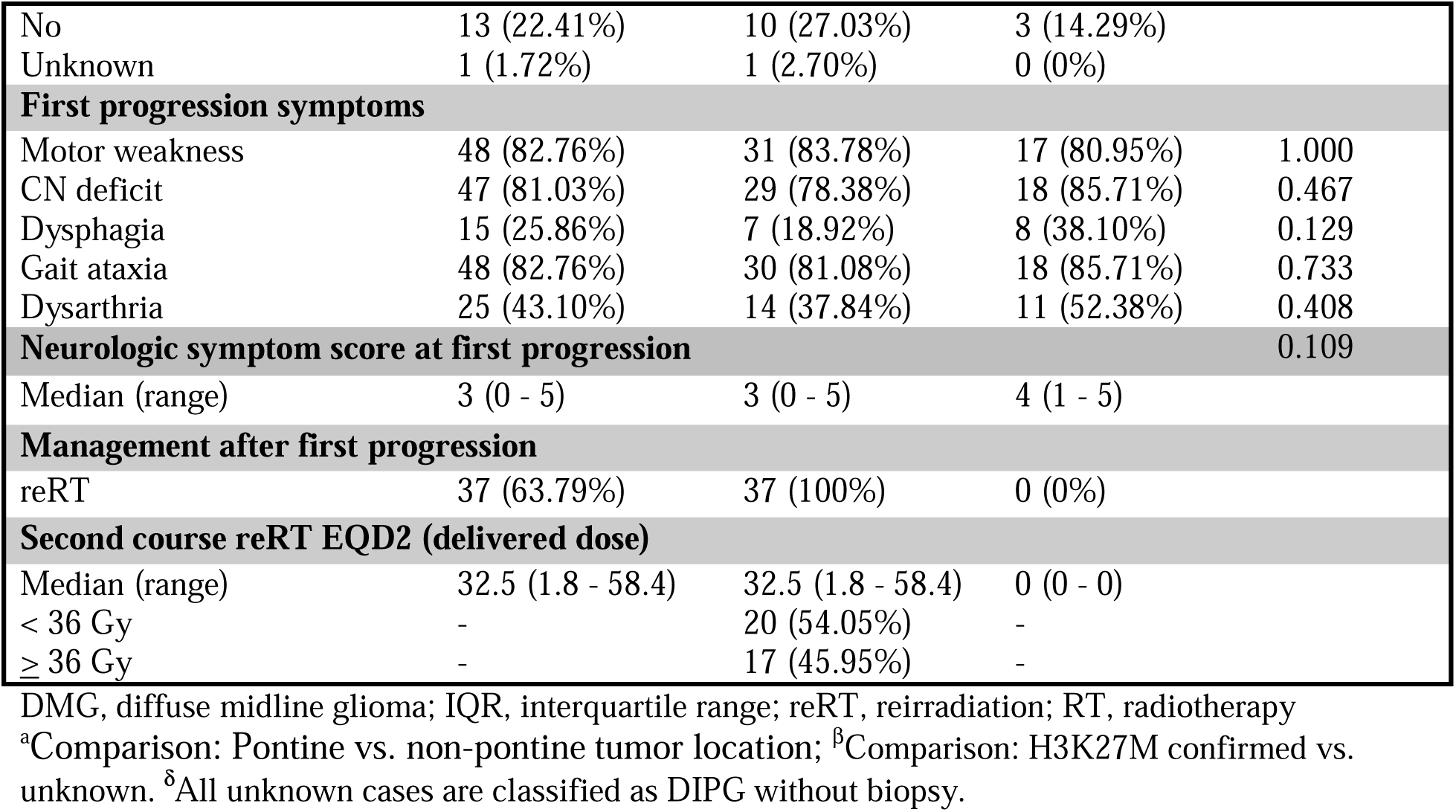
Patient characteristics.

| Characteristics | Overall (%)<br>N = 58 | reRT (%)<br>N = 37 | No reRT (%)<br>N = 21 | P-value |
| --- | --- | --- | --- | --- |
| **Sex** |  |  |  | 0.786 |
| Female | 28 (48.28%) | 17 (45.95%) | 11 (52.38%) |  |
| Male | 30 (51.72%) | 20 (54.05%) | 10 (47.62%) |  |
| **Age at diagnosis [years]** |  |  |  | 0.962 |
| Median (range) | 7.7 (3.2 - 26.7) | 7.4 (3.2 - 26.7) | 7.8 (4.2 - 16.9) |  |
| **Diagnosis year** |  |  |  | 0.035 |
| 2011-2017 | 17 (29.31%) | 7 (18.92%) | 10 (47.62%) |  |
| 2018-2025 | 41 (70.69%) | 30 (81.08%) | 11 (52.38%) |  |
| **Tumor location** |  |  |  | 0.347<sup>α</sup> |
| Pons | 43 (74.14%) | 25 (67.57%) | 18 (85.71%) |  |
| Thalamus | 13 (22.41%) | 10 (27.03%) | 3 (14.29%) |  |
| Midbrain | 2 (3.45%) | 2 (5.41%) | 0 (0%) |  |
| **Molecular confirmation of DMG** |  |  |  | 0.750<sup>β</sup> |
| H3K27M | 46 (79.31%) | 29 (78.38%) | 17 (80.95%) |  |
| EZHIP overexpression | 2 (3.44%) | 2 (5.41%) | 0 (0%) |  |
| Unknown<sup>δ</sup> | 10 (17.25%) | 6 (16.22%) | 4 (19.05%) |  |
| **Extent of resection** |  |  |  | 0.373 |
| No biopsy | 10 (17.24%) | 5 (13.51%) | 5 (23.81%) |  |
| Biopsy | 38 (65.52%) | 24 (64.86%) | 14 (66.67%) |  |
| Subtotal resection | 10 (17.24%) | 8 (21.62%) | 2 (9.52%) |  |
| **Required steroids on presentation** |  |  |  | 0.076 |
| Yes | 41 (70.69%) | 23 (62.16%) | 18 (85.71%) |  |
| No | 17 (29.31%) | 14 (37.84%) | 3 (14.29%) |  |
| **Presenting symptoms** |  |  |  |  |
| Motor weakness | 43 (74.14%) | 29 (78.38%) | 14 (66.67%) | 0.363 |
| CN deficit | 42 (72.41%) | 25 (67.57%) | 17 (80.95%) | 0.365 |
| Dysphagia | 16 (27.59%) | 10 (27.03%) | 6 (28.57%) | 1.000 |
| Gait Ataxia | 47 (81.03%) | 28 (75.68%) | 19 (90.48%) | 0.296 |
| Dysarthria | 19 (32.76%) | 12 (32.43%) | 7 (33.33%) | 1.000 |
| **Neurologic symptom score at presentation** |  |  |  | 0.674 |
| Median (range) | 3 (0 - 5) | 3 (0 - 5) | 3(1 - 4) |  |
| **First course RT (delivered dose)** |  |  |  | 0.681 |
| Median dose [Gy] (range) | 54 (39 - 60) | 54 (39 - 60) | 54 (39 - 59.4) |  |
| Hypo-fractionation | 2 (3.45%) | 1 (2.70%) | 1 (4.76%) |  |
| Conventional fractionation | 56 (96.55%) | 36 (97.30%) | 20 (95.24%) |  |
| **Progression site** |  |  |  | 0.164 |
| Brain Local | 44 (75.86%) | 27 (72.97%) | 17 (80.95%) |  |
| Brain Distant | 5 (8.62%) | 2 (5.41%) | 3 (14.29%) |  |
| Brain & Spinal Cord | 9 (15.52%) | 8 (21.62%) | 1 (4.76%) |  |
| **Median time from diagnosis to first progression [months]** |  |  |  | 0.067 |
| Median (IQR) | 8.7 (6.43 - 11.7) | 10.1 (6.9 - 11.8) | 6.9 (5.1 - 10.6) |  |
| **Required steroids at first progression** |  |  |  | 0.333 |
| Yes | 44 (75.86%) | 26 (70.27%) | 18 (85.71%) |  |
| No | 13 (22.41%) | 10 (27.03%) | 3 (14.29%) |  |
| Unknown | 1 (1.72%) | 1 (2.70%) | 0 (0%) |  |
| **First progression symptoms** |  |  |  |  |
| Motor weakness | 48 (82.76%) | 31 (83.78%) | 17 (80.95%) | 1.000 |
| CN deficit | 47 (81.03%) | 29 (78.38%) | 18 (85.71%) | 0.467 |
| Dysphagia | 15 (25.86%) | 7 (18.92%) | 8 (38.10%) | 0.129 |
| Gait ataxia | 48 (82.76%) | 30 (81.08%) | 18 (85.71%) | 0.733 |
| Dysarthria | 25 (43.10%) | 14 (37.84%) | 11 (52.38%) | 0.408 |
| **Neurologic symptom score at first progression** |  |  |  | 0.109 |
| Median (range) | 3 (0 - 5) | 3 (0 - 5) | 4 (1 - 5) |  |
| **Management after first progression** |  |  |  |  |
| reRT | 37 (63.79%) | 37 (100%) | 0 (0%) |  |
| **Second course reRT EQD2 (delivered dose)** |  |  |  |  |
| Median (range) | 32.5 (1.8 - 58.4) | 32.5 (1.8 - 58.4) | 0 (0 - 0) |  |
| < 36 Gy | - | 20 (54.05%) | - |  |
| ≥ 36 Gy | - | 17 (45.95%) | - |  |

DMG, diffuse midline glioma; IQR, interquartile range; reRT, reirradiation; RT, radiotherapy
<sup>a</sup>Comparison: Pontine vs. non-pontine tumor location; <sup>β</sup>Comparison: H3K27M confirmed vs. unknown. <sup>δ</sup>All unknown cases are classified as DIPG without biopsy.

### Pattern of failure at first progression

The median time from diagnosis to first disease progression was 8.70 months (IQR: 6.43–11.7 months). There was a trend towards longer time from diagnosis to first disease progression for patients treated with reRT (10.07 vs. 6.87 months, p=0.066). Most common site of failure was local within the site of the primary tumor (75.86%). Distant failures were represented by progression in a site of the brain that was distinct from the primary (8.62%) or combined progression within the brain and spinal cord, often with leptomeningeal involvement (15.52%).

### Neurologic symptoms

Similar to initial presentation, motor weakness (82.76%), cranial nerve deficits (81.03%), and gait ataxia (82.76%) were the predominant neurologic symptoms at the time of first progression. The composite neurologic symptom score at first progression did not significantly differ in the no reRT vs. reRT group (median score: 4 vs 3, p=0.109). Most patients (75.86%) required steroids at first progression, and this did not differ by treatment group (p= 0.333). Five patients (n=3 from reRT; n=2 from no reRT) were missing neurologic symptom data after second-line treatment. Following second-line treatment, the reRT vs. no reRT group demonstrated greater improvement in motor function (51.4% vs. 9.5%, p=0.002), cranial nerve function (29.7% vs. 4.8%, p=0.044), and gait ataxia (35.1% vs. 9.5%, p=0.059) (**Figure 1**). The effect sizes of improvement in dysarthria (8.1% vs. 0%, p=0.504) and dysphagia (10.8% vs. 4.8%, p=0.796) were small and not statistically significant.

**Figure 1.**
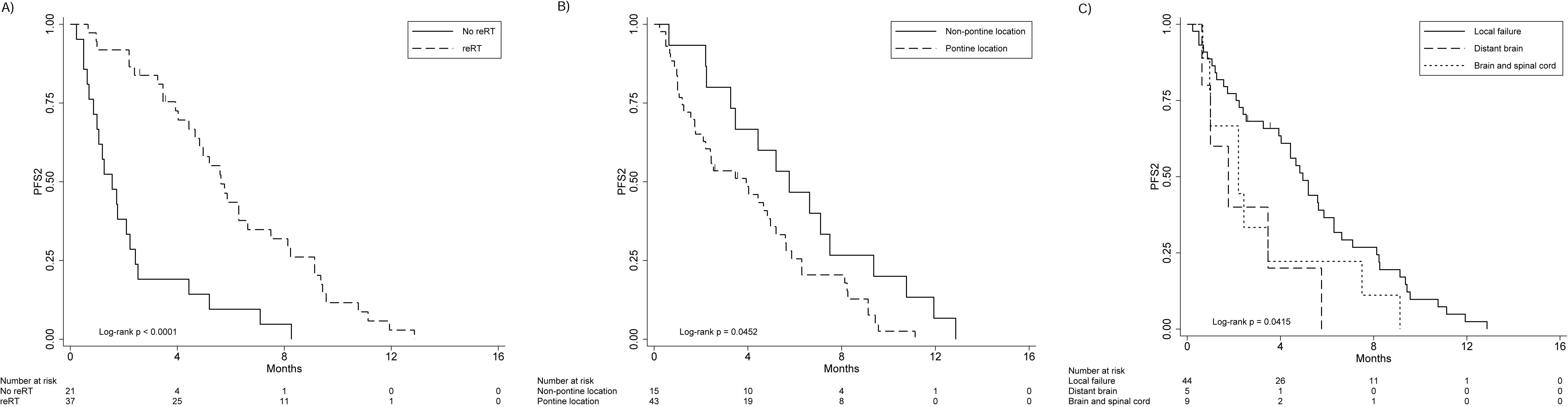
Neurological symptoms after second-line treatment in the reRT and no reRT groups. *Comparison of improved symptoms between the reRT and no reRT groups (p < 0.05). Five patients have missing data (n = 3 in the reRT group and n = 2 in the no reRT group). ** Comparison of improved symptoms between the reRT and no reRT groups (p < 0.01). Five patients have missing data (n = 3 in the reRT group and n = 2 in the no reRT group).

A greater proportion of patients in the no reRT vs. reRT group demonstrated worsening of motor function (52.4.% vs. 16.2%, p=0.009), cranial nerve function (52.4.% vs. 13.5%, p=0.004) and gait ataxia (57.1% vs. 16.2% p=0.004).

### Radiation at first progression

Among patients who underwent reRT, the median delivered equivalent of 2 Gy dose (EQD2) was 32.5 Gy (range: 1.8-58.4 Gy) and 45.9% of patients received an EQD2 ≥ 36 Gy. The median time from first progression to reRT start was 28 days (IQR: 19–50 days). All but two patients (94.6%) completed their prescribed dose of reRT. The two patients who did not complete reRT presented with intractable seizures were found to have leptomeningeal disease in the brain and spine and developed rapid symptomatic disease progression. Both patients elected for supportive care after the first fraction of reRT. For local failures and distant brain failures (n=29), reRT fields focally encompassed progressive disease in the brain. For distant failures involving both the brain and spine (n=8), reRT field included the craniospinal volume except for one patient.

This patient’s treatment volume included the largest region of gross disease in the cervical spine, and this was one of the patients who only completed one fraction of reRT before deciding to pursue supportive care.

### Overall survival

The median OS1 of the entire cohort was 15.83 months (95% Confidence Interval [CI]: 12.77–20.17 months) and median OS2 was 5.6 months (95% CI: 5.03–9.67 months). The median OS1 was 20.17 months (95% CI: 14.00–23.03 months) with reRT and 11.07 months (95% CI: 7.77–15.83 months) without reRT. The median OS2 was 9.67 months (95% CI: 5.60–11.97 months) with reRT and 2.57 months (95% CI: 1.77–5.57 months) without reRT. ReRT was associated with improved OS2 (6-month OS2: 64.5% vs. 14.3%, 95% CI: 45.9%–78.2% vs. 3.6%–32.1%; log-rank p<0.001) (**Figure 2A**). The following variables were also associated with a worse OS2 in the entire cohort: pontine vs. non-pontine tumor location (6-month OS2: 34.5% vs. 77.8%, 95% CI: 20.9%–49.4% vs. 45.5%–92.3%; log-rank p=0.0054) (**Figure 2B**) and use of steroids at first progression (6-month OS2: 35.% vs. 75.5%, 95% CI: 21.4%–50.1% vs. 41.6%–91.4%; log-rank p=.0066) (**Figure 2C**). Steroid use at first progression was associated with a higher burden of neurologic symptoms (median sum 4 vs. 2, p = 0.005).

**Figure 2.**
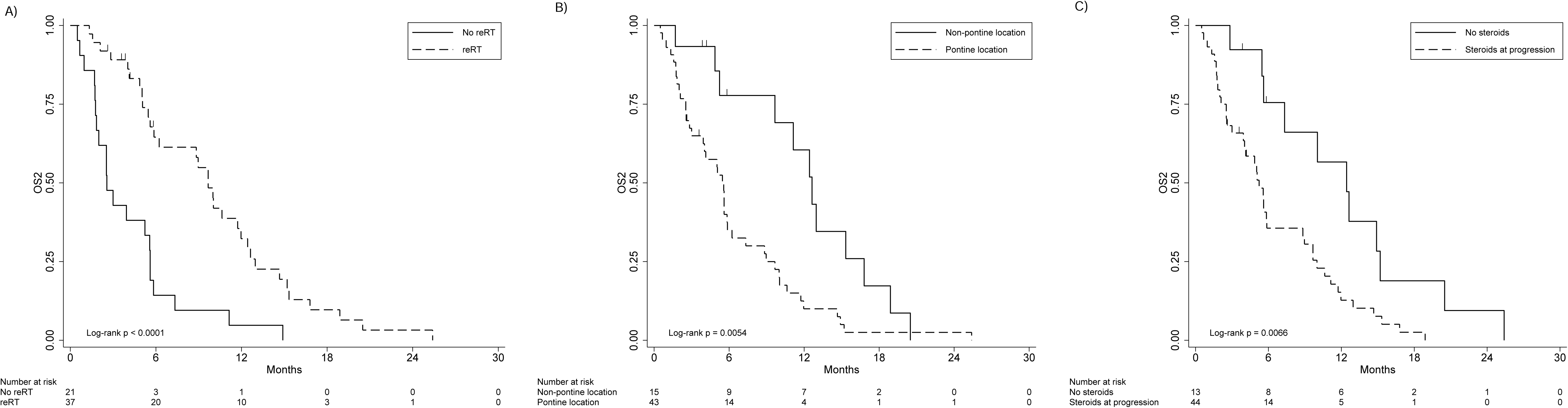
Overall survival as measured from first progression (OS2) by A) re-RT, B) tumor location, and C) receipt of steroids

On univariable analysis, OS2 was associated with reRT, time to first progression from diagnosis, composite neurologic symptom score at progression, use of steroids at first progression and pontine vs. non-pontine tumor location (**Table 2**). On multivariable analysis, reRT remained independently associated with improved OS2 (HR: 0.27, 95% CI: 0.14–0.51, p <0.0001). Pontine vs. non-pontine tumor location (HR: 2.94, 95% CI: 1.40–6.15, p=.004) and use of steroids at first progression (HR: 4.12, 95% CI:1.83–9.29, p =.001) remained independently associated with worse OS2. OS2 was not associated with sex, extent of resection, or reRT EQD2.

**Table 2.**
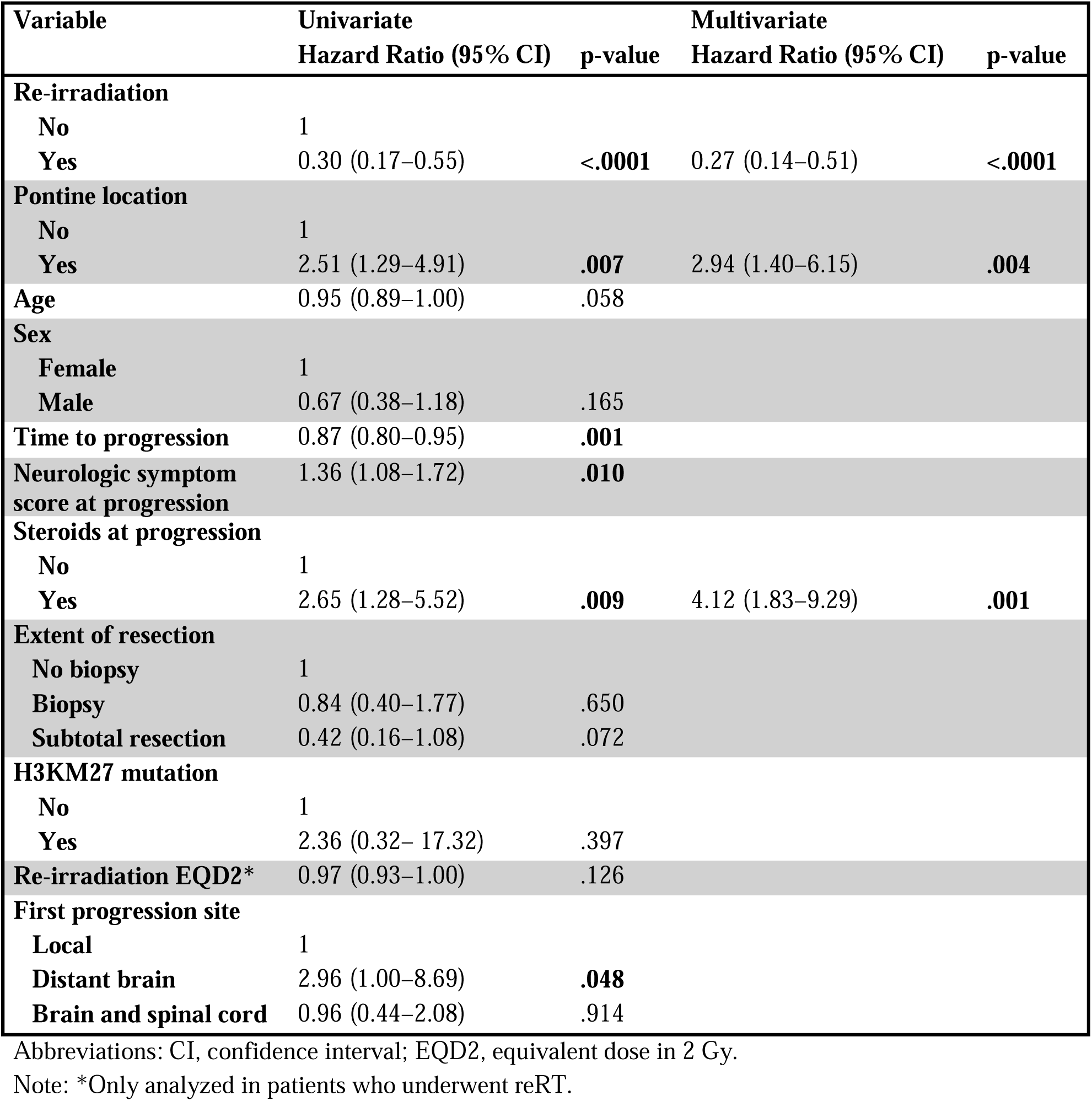
Univariate and multivariable Cox regression analyses evaluating predictors of OS2.

| **Variable** | **Univariate**<br>**Hazard Ratio (95 % CI)** | **p-value** | **Multivariate**<br>**Hazard Ratio (95 % CI)** | **p-value** |
| --- | --- | --- | --- | --- |
| **Re-irradiation** |  |  |  |  |
| **No** | 1 |  |  |  |
| **Yes** | 0.30 (0.17–0.55) | **<.0001** | 0.27 (0.14–0.51) | **<.0001** |
| **Pontine location** |  |  |  |  |
| **No** | 1 |  |  |  |
| **Yes** | 2.51 (1.29–4.91) | **.007** | 2.94 (1.40–6.15) | **.004** |
| **Age** | 0.95 (0.89–1.00) | .058 |  |  |
| **Sex** |  |  |  |  |
| **Female** | 1 |  |  |  |
| **Male** | 0.67 (0.38–1.18) | .165 |  |  |
| **Time to progression** | 0.87 (0.80–0.95) | **.001** |  |  |
| **Neurologic symptom score at progression** | 1.36 (1.08–1.72) | **.010** |  |  |
| **Steroids at progression** |  |  |  |  |
| **No** | 1 |  |  |  |
| **Yes** | 2.65 (1.28–5.52) | **.009** | 4.12 (1.83–9.29) | **.001** |
| **Extent of resection** |  |  |  |  |
| **No biopsy** | 1 |  |  |  |
| **Biopsy** | 0.84 (0.40–1.77) | .650 |  |  |
| **Subtotal resection** | 0.42 (0.16–1.08) | .072 |  |  |
| **H3KM27 mutation** |  |  |  |  |
| **No** | 1 |  |  |  |
| **Yes** | 2.36 (0.32– 17.32) | .397 |  |  |
| **Re-irradiation EQD2\*** | 0.97 (0.93–1.00) | .126 |  |  |
| **First progression site** |  |  |  |  |
| **Local** | 1 |  |  |  |
| **Distant brain** | 2.96 (1.00–8.69) | **.048** |  |  |
| **Brain and spinal cord** | 0.96 (0.44–2.08) | .914 |  |  |

Abbreviations: CI, confidence interval; EQD2, equivalent dose in 2 Gy.
Note: \*Only analyzed in patients who underwent reRT.

On landmark analysis, after exclusion of patients who died within one month of progression (n=3), the median OS2 for patients who received reRT was 9.67 months (95% CI: 5.60–11.97 months) compared to 3.0 months (95% CI: 2.0–5.60 months) without reRT (log-rank p<0.001) (**Supplemental Figure 2**). After adjusting for confounding variables, ReRT remained independently associated with improved OS2 (HR: 0.28, 95% CI: 0.14–0.55, p <0.0001). Pontine versus non-pontine tumor location, use of steroids at first progression, and distant pattern of failure were also independently associated with worse OS2 (**Supplemental Table 1**).

In the as-treated analysis, the two patients with leptomeningeal disease who started reRT but chose not to complete therapy after the first fraction were removed. As expected, reRT remained associated with improved OS2 when these patients were removed (6-month OS2: 68.2% vs. 14.3%; 95% CI: 48.9%–81.5% vs. 3.6%–32.1%; log-rank p <0.001).

### Progression-free survival

The median PFS1 of the entire cohort was 8.53 months (95% CI: 7.07–10.57 months) and median PFS2 was 4.43 months (95% CI: 2.43–5.60 months). The median PFS1 was 6.87 months (95% CI, 5.13–10.5 months) without reRT and 10.07 months (95% CI: 7.47–11.2 months) with reRT. The median PFS2 without reRT was 1.57 months (95% CI: 0.87–2.23 months) and 5.63 months with reRT (95% CI: 4.43–7.50 months). ReRT was associated with improved PFS2 (6-month PFS2: 43.5% vs. 9.5%, 95% CI: 26.9%–58.9% vs. 1.6%–26.1%; log-rank p <0.0001)

(**Figure 3A**). Worse PFS2 was associated with pontine vs. non-pontine tumor location (6-month PFS2: 25.5% vs. 46.7%, 95% CI: 13.4%–39.5% vs. 21.2%–68.8%; log-rank p=0.0452) (**Figure 3B**) and progression site (3-month PFS2: local failure 68.2%, 95% CI: 52.3%–79.8%; distant brain 40%, 95% CI: 5.2%–75.3%; distant spine and brain 33.3%, 7.8%–62.3%; log-rank p=.0415) (**Figure 3C**).

**Figure 3.**
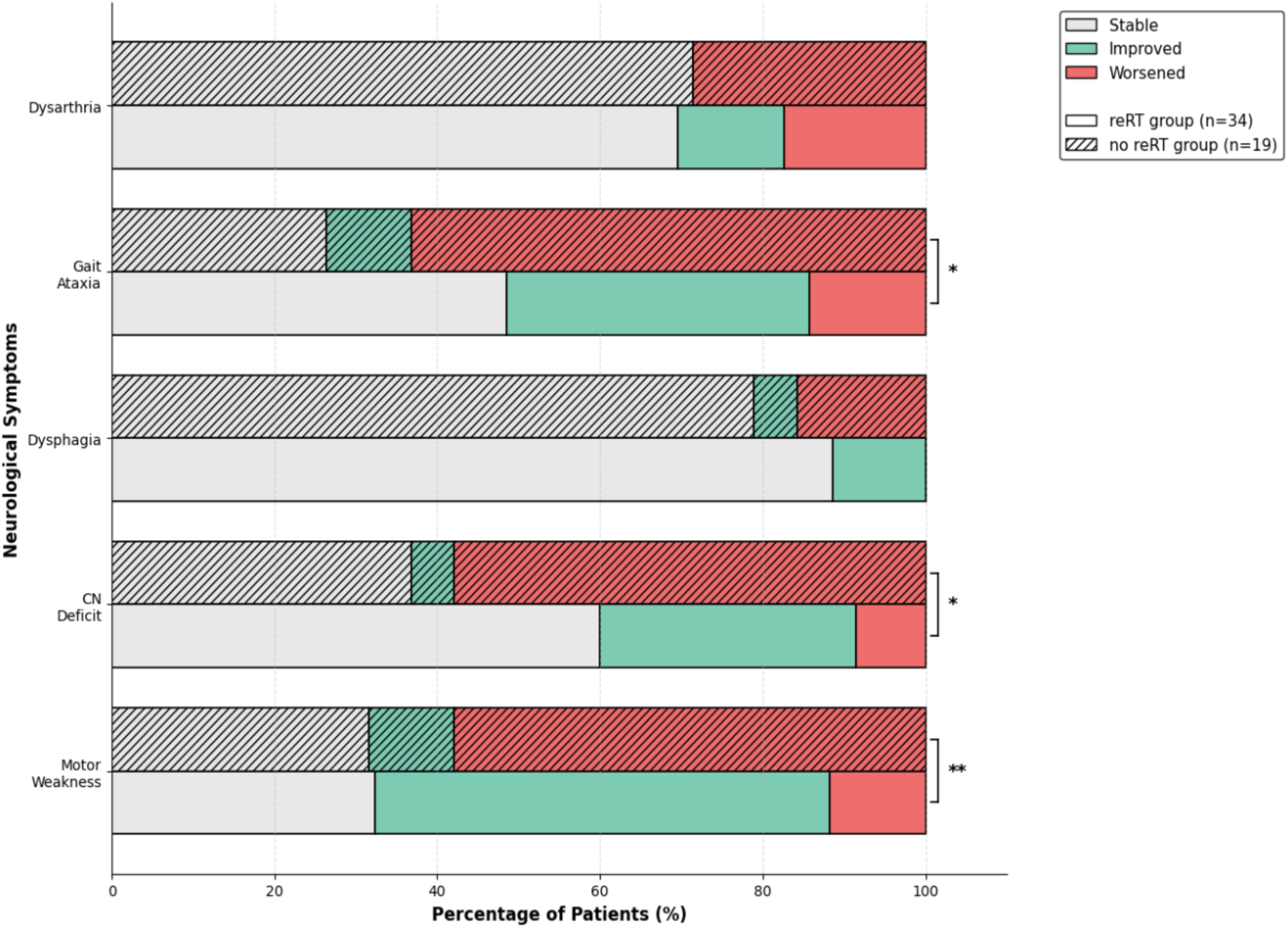
Progression free survival as measured from first progression (PFS2) by A) re-RT, B) tumor location, and C) site of failure

On univariable analysis, PFS2 was associated with reRT, pontine location, time to first progression from diagnosis, and distant brain progression (Table 3). On multivariable analysis, reRT remained independently associated with improved PFS2 (HR: 0.23, 95% CI: 0.12–0.42, p <0.0001). Distant pattern of first failure was independently associated with worsened PFS2 (distant brain vs. local failure: HR 2.83, 95% CI 1.07–7.51, p=.037; distant spine and brain vs. local failure: HR 2.49, 95% CI 1.17–5.30, p=.018). PFS2 was not associated with sex, extent of resection, or reRT EQD2.

**Table 3.** Univariate and multivariable Cox regression analyses evaluating predictors of PFS2.

| **Variable** | **Univariate**<br>**Hazard ratio (95% CI)** | **p-value** | **Multivariate**<br>**Hazard ratio (95% CI)** | **p-value** |
| --- | --- | --- | --- | --- |
| **Re-irradiation** |  |  |  |  |
| **No** | 1 |  | 1 |  |
| **Yes** | 0.26 (0.14–0.47) | **<.0001** | 0.23 (0.12–0.42) | **<.0001** |
| **Pontine location** |  |  |  |  |
| **No** | 1 |  |  |  |
| **Yes** | 1.89 (0.99–3.57) | **.050** |  |  |
| **Age** | 0.99 (0.94–1.04) | .673 |  |  |
| **Sex** |  |  |  |  |
| **Female** | 1 |  |  |  |
| **Male** | 0.92 (0.54–1.57) | .770 |  |  |
| **Time to progression** | 0.95 (0.90–0.99) | **.049** |  |  |
| **Neurologic symptom score at progression** | 1.10 (0.89–1.36) | .368 |  |  |
| **Steroids at progression** |  |  |  |  |
| **No** | 1 |  |  |  |
| **Yes** | 1.62 (0.85–3.07) | .141 |  |  |
| **Extent of resection** |  |  |  |  |
| **No biopsy** | 1 |  |  |  |
| **Biopsy** | 1.03 (0.49–2.15) | .934 |  |  |
| **Subtotal resection** | 0.60 (0.24–1.54) | .291 |  |  |
| **H3KM27 mutation** |  |  |  |  |
| **No** | 1 |  |  |  |
| **Yes** | 1.12 (0.27– 4.67) | .879 |  |  |
| **Re-irradiation EQD2\*** | 0.96 (0.93–1.00) | .051 |  |  |
| **First progression site** |  |  |  |  |
| **Local** | 1 |  |  |  |
| **Distant brain** | 2.70 (1.04–7.04) | **.042** | 2.83 (1.07–7.51) | **.037** |
| **Brain and spinal cord** | 1.87 (0.89–3.92) | .095 | 2.49 (1.17–5.30) | **.018** |

Abbreviations: CI, confidence interval; EQD2, equivalent dose in 2 Gy.
Note: \*Only analyzed in patients who underwent reRT.

By landmark analysis, the median PFS2 for patients who received reRT was 5.63 months (95% CI: 4.43–7.50 months) compared to 1.73 months (95% CI: 1.07–2.43 months) without reRT (log-rank p<.0001) **(Supplemental Figure 3)**. On multivariable analysis, reRT remained independently associated with improved PFS2 (HR: 0.25, 95% CI: 0.13–0.47, p < 0.0001) as well as site of progression (**Supplemental Table 2**).

### Factors predictive of survival in the reRT treatment group

Given that tumor location, steroid use and pattern of first failure were prognostic in our study population, we sought to determine whether these factors were also predictive of better OS2 and PFS2 after reRT. Within the reRT treatment group (n=37), non-pontine vs. pontine tumor location was associated with a doubling of median OS2 (6.23 vs. 12.97 months, 95% CI: 5.03–10 vs. 4.87–18.9 months, log-rank p=0.020), highlighting that patients with non-pontine tumors derived a greater benefit from reRT (**Supplemental Figure 4**). Use of steroids and pattern of first failure (distant vs. local) was not predictive of OS2 after reRT (all log-rank p>0.05). With respect to PFS2, distant failure was predictive of worse outcomes after reRT (median PFS2: 2.20 vs. 6.30 months, 95% CI: 0.67–5.77 vs. 4.97–9.13 months, log-rank p=0.0029), suggesting that pattern of failure was predictive of PFS2 but not OS2 (**Supplemental Figure 5**). Neither tumor location nor use of steroids was predictive of PFS2 after reRT (all log-rank p>0.05).

### Dosimetry

Among patients who underwent reRT, organ-at-risk (OAR) dosimetric parameters were available in most patients. Median (range) maximum EQD2 doses were 68.08 Gy (range: 33.09–78.44 Gy) to the optic apparatus (n=27), 88.27 Gy (range: 68.02–100.01 Gy) to the uninvolved brainstem (n=19), 55.10 Gy (range: 8.73–64.27 Gy) and 54.88 Gy (range: 20.63–74.96 Gy) to the left and right cochleae, respectively, (each n=26), and 73.24 Gy (range: 2.33–92.47 Gy) to the spinal cord (n=21) (**Supplemental Table 3**).

## Discussion

Treatment with reRT at first progression was associated with improvement in neurologic symptoms as well as a significant OS2 and PFS2 benefit. These findings support reRT as one of the few available interventions capable of achieving symptomatic stabilization and modest extension of survival in the post-progression setting. Importantly, these findings arise from a contemporary cohort in which the majority of patients had molecularly characterized DMG, reflecting current diagnostic standards and extending beyond historical pontine DMG-only series. Patients diagnosed between 2018 and 2025 were more likely to undergo reRT compared with those diagnosed earlier (81.1% vs. 52.4%, p =.035), suggesting increasing adoption of reRT. This cohort also includes a substantial proportion of non-pontine DMG, a population for which outcome data particularly in the reRT setting remain limited. We demonstrate here that among the patients who were treated with reRT, those with non-pontine tumors, most commonly located in the thalamus, and those with local failures derived the greatest benefit in terms of OS2 and PFS2, respectively.

In our cohort, the median OS2 and PFS2 with reRT was 5.6 months and 4.43 months, respectively. ReRT conferred a substantial OS2 advantage, with median survival from first progression nearly tripling compared with patients who did not receive reRT. After adjusting for relevant clinical and molecular variables, reRT remained independently associated with improved OS2 (HR 0.27, 95% CI: 0.14–0.51). These results are consistent with prior retrospective series in which reRT consistently prolonged post-relapse survival with reported median OS2 ranging from 4 to 8 months.^18,19,25–27^ Shariff et al. recently summarized survival outcomes across these studies.^19^ Notably, most prior reports lacked a comparison cohort of patients that did not receive a second course of RT and rarely integrated neurologic or molecular data, limiting interpretation of treatment effect and prognostic implications.

Time from diagnosis to progression emerged as a prognostic indicator of OS2 and PFS2 on univariable analysis; however, it did not retain independent association on multivariable modeling. This conflicts with prior literature.^19^ In a matched-cohort analysis by the SIOP-E-HGG/DIPG working group, interval to progression was independently prognostic of survival.^15^ Similarly, in the aforementioned study by Shariff et al., patients with a latent interval >1 year between initial RT and reRT experienced substantially longer OS after reRT (median 10.9 vs. 5.5 months, p=0.023).^19^ Beyond clinical timing of progression, the impact of radiographic progression features is also being explored. Vajapeyam et al. demonstrated that elevated relative cerebral blood volume on perfusion MRI predicted shorter PFS and OS.^28^ Taken together, these findings suggest that both temporal and radiographic features of progression reflect the biologic heterogeneity in DMG and may impact responsiveness to salvage therapy. The lack of independent prognostic significance in our cohort may reflect sample size limitations, particularly due to the relatively small number of patients that progressed after 12 months (n =13).

The classic presentation of pontine DMG includes cranial nerve deficits, long tract signs, and cerebellar dysfunction.^29^ The predominant symptoms of cranial nerve palsies, ataxia, and motor weakness observed in greater than two-thirds of our cohort closely mirror patterns described in prior epidemiologic patterns.^30^A strength of our study is the characterization of neurologic symptoms at first progression and after second line therapy, which directly reflects patient quality of life yet is often underreported. Patients who underwent reRT demonstrated significantly greater improvement in motor function (p =0.002), cranial nerve function (p=0.037), and gait ataxia (p=0.046). In contrast, patients who did not receive reRT were substantially more likely to have worsened across these same three neurological domains. Because patients with progressive DIPG/DMG often experience rapid neurologic decline, palliation is critical, and corticosteroids have remained central to symptom management. Notably, steroid use at progression was independently associated with worse OS2 in our cohort, highlighting the interplay of symptomatic burden, steroid dependence, and survival. Patients that received reRT demonstrated less neurologic decline at follow up compared with those who did not suggesting a potential steroid-sparing benefit. Prospective evaluation of corticosteroid trajectories in the context of reRT is warranted.

In our cohort, pontine tumor location was found to be strongly associated with worse OS2 as well as predictive of OS2 within the reRT treatment group. These findings are consistent with prior studies demonstrating that DMGs arising in non-brainstem midline regions, such as the thalamus and spinal cord, tend to occur in older patients and are associated with better OS.^31,32^ In a cohort of 47 patients with *H3 p.K27M*-mutant DMGs, patients with pontine tumors were younger (median age 7 years) compared to those with thalamic (median 24 years) or spinal (median 25 years) tumors.^33^ Interestingly, in a study by Wang et al. evaluating 121 DMGs across multiple anatomic locations, *H3 p.K27M* mutation status was strongly associated with poorer prognosis in infratentorial gliomas, such as the brainstem, whereas the mutation conferred no significant prognostic impact in supratentorial gliomas such as those arising in the thalamus.^32^ A large systematic review of 804 *H3 p.K27M*-mutant DMGs found that patients with brainstem tumors had significantly worse survival compared to those with thalamic (HR 0.57, 95% CI 0.46-0.71; p < 0.001) or spinal cord lesions (HR 0.46, 95% CI 0.34-0.62; p < 0.001).^31^ These findings reinforce brainstem involvement as a dominant adverse prognostic factor. Tumors arising here represent a distinct subgroup, characterized by earlier age at presentation, more aggressive disease behavior, with outcomes further constrained by critical neural structures and limited surgical feasibility inherent to the brainstem.

Similar to prior studies, we observed no dose-response relationship with reRT.^26^ Dose and fractionation widely varied in our cohort with both hypofractionation and conventional fractionation approaches used. Recent studies have demonstrated comparable survival outcomes between hypofractionation and conventional approaches.^27^ Prospective data from the small phase I/II trial by Amsbaugh et al. demonstrated no clinical benefit with modest dose escalation beyond 24 Gy in 12 fractions for progressive DIPG.^34^ A prospective single-arm phase II trial evaluating 30.6 Gy or 36 Gy by conventional fractionation will provide further evidence regarding the dosage with reRT (NCT03126266). NCCN guidelines acknowledge hypofractionation as an emerging alternative; however, the optimal reRT dose and fractionation schedule remains undefined.^35^ In this context, regimen selection should be individualized, incorporating time since prior RT, performance status, symptom trajectory, and goals of care.

Limitations of this study include retrospective nature which limits generalizability. Lack of standardized neurological symptom or quality of life assessment tools preclude in-depth analysis, and there is an unaddressed need to validate these instruments in DIPG/DMG. There may be potential selection bias in favor of reRT, although our study found that median time to progression after initial RT was similar between the two groups, reducing an aspect of this concern. Finally, landmark analyses were performed to mitigate immortal time bias and improve reliability of survival estimates.

In conclusion, in patients with DIPG/DMG, reRT was associated with neurologic improvement and a moderate prolongation of overall survival of seven months from first progression. Patients with non-pontine tumors and local failure may derive the greatest benefit. Prospective investigation is needed to establish optimal dose/fractionation and to identify prognostic factors across both pontine and non-pontine tumor locations to help guide patient selection.

## Ethics Approval Statement

Approval was granted by the Johns Hopkins Institutional Review Board (IRB No. IRB00324381). Given this is a retrospective study, informed consent was waived.

## Author Contributions

SA and ML conceived the study. TV and DV performed data collection. TV and SA performed statistical analysis. TV, DV, and SA drafted the manuscript. MA, CHL, MG, LK, EH, RP, RDG, MAK, ER, KC, and ML edited the manuscript. All authors approved the final manuscript.

## Data Availability

Data is available upon request to the corresponding author and approval by the Institution’s Review Board.

