## Supplemental Tables 1-3, Supplemental Figures 1-5 for "Survival and neurologic outcomes after re-irradiation in children with diffuse midline glioma and diffuse intrinsic pontine glioma"

**Supplemental Table 1.** Univariate and multivariable Cox regression analyses evaluating predictors of OS2 on landmark analysis

| **Prognostic** | **Univariate** | | **Multivariate** | |
|---|---|---|---|---|
| | **Hazard ratio (95% CI)** | **p-value** | **Hazard ratio (95% CI)** | **p-value** |
| **Re-irradiation** | | | | |
| **No** | 1 | | | |
| **Yes** | 0.33 (0.18–0.62) | **.001** | 0.28 (0.14–0.55) | **<.0001** |
| **Pontine location** | | | | |
| **No** | 1 | | | |
| **Yes** | 2.40 (1.22–4.72) | **.011** | 3.13 (1.41–6.95) | **.005** |
| **Age** | 0.95 (0.90–1.00) | .073 | | |
| **Sex** | | | | |
| **Female** | 1 | | | |
| **Male** | 0.68 (0.38–1.23) | .206 | | |
| **Time to progression** | 0.89 (0.82–0.96) | **.005** | | |
| **Composite neurologic symptoms at progression** | 1.32 (1.04–1.67) | **.021** | | |
| **Steroids at progression** | 2.54 (1.21–5.31) | **.013** | 3.72 (1.65–8.40) | **.002** |
| **Extent of resection** | | | | |
| **No biopsy** | 1 | | | |
| **Biopsy** | 0.76 (0.36–1.60) | .464 | | |
| **Subtotal resection** | 0.40 (0.16–1.03) | .058 | | |
| **H3KM27 mutation** | | | | |
| **No** | 1 | | | |
| **Yes** | 2.23 (0.30– 16.37) | .431 | | |
| **Re-irradiation EQD2** | 0.97 (0.93–1.00) | .126 | | |
| **First progression site** | | | | |
| **Local** | | | | |
| **Distant brain** | 3.72 (1.25–11.06) | **.018** | 5.97 (1.78–19.96) | **.004** |
| **Brain and spinal cord** | 1.04 (0.47–2.27) | .928 | | |

Abbreviations: CI, confidence interval; EQD2, equivalent dose in 2 Gy.

**Supplemental Table 2.** Univariate and multivariable Cox regression analyses evaluating predictors of PFS2 on landmark analysis

| **Variable** | **Univariate**<br>**Hazard ratio (95% CI)** | **p-value** | **Multivariate**<br>**Hazard ratio (95% CI)** | **p-value** |
|---|---|---|---|---|
| **Re-irradiation** | | | | |
| **No** | 1 | | 1 | |
| **Yes** | 0.28 (0.15–0.53) | **<.0001** | 0.25 (0.13–0.47) | **<.0001** |
| **Pontine location** | | | | |
| **No** | 1 | | | |
| **Yes** | 1.80 (0.95–3.42) | .074 | | |
| **Age** | 0.99 (0.94–1.0) | .776 | | |
| **Sex** | | | | |
| **Female** | 1 | | | |
| **Male** | 0.95 (0.55–1.65) | .868 | | |
| **Time to progression** | 0.96 (0.9–1.01) | .097 | | |
| **Neurologic symptom score at progression** | 1.07 (0.86–1.32) | .563 | | |
| **Steroids at progression** | 1.54 (0.80–2.93) | .194 | | |
| **Extent of resection** | | | | |
| **No biopsy** | 1 | | | |
| **Biopsy** | 0.94 (0.45–1.98) | .877 | | |
| **Subtotal resection** | 0.59 (0.23–1.50) | .266 | | |
| **H3KM27 mutation** | | | | |
| **No** | 1 | | | |
| **Yes** | 1.10 (0.26– 4.61) | .894 | | |
| **Re-irradiation EQD2** | 0.96 (0.93–1.00) | **.051** | | |
| **Progression Site** | | | | |
| **Local** | 1 | | 1 | |
| **Distant brain** | 3.03 (1.15–7.93) | **.020** | 3.42 (1.28–9.17) | **.015** |
| **Brain and spinal cord** | 2.10 (0.99–4.43) | **.050** | 2.70 (1.26–5.80) | **.011** |

Abbreviations: CI, confidence interval; EQD2, equivalent dose in 2 Gy.

**Supplemental Table 3.** Organ-at-risk dosimetric parameters in patients that underwent reirradiation (N=37).

| Organ-at-risk | Median Maximum EQD2 (range) |
|---|---|
| **Optic apparatus (n=27)** | 68.08 (33.09–78.44) |
| **Brainstem (n=19)** | 88.27 (68.02–100.01) |
| **Left Cochlea (n=26)** | 55.10 (8.73–64.27) |
| **Right Cochlea (n=26)** | 54.88 (20.63–74.96) |
| **Spinal Cord (n=21)** | 73.24 (2.33–92.47) |

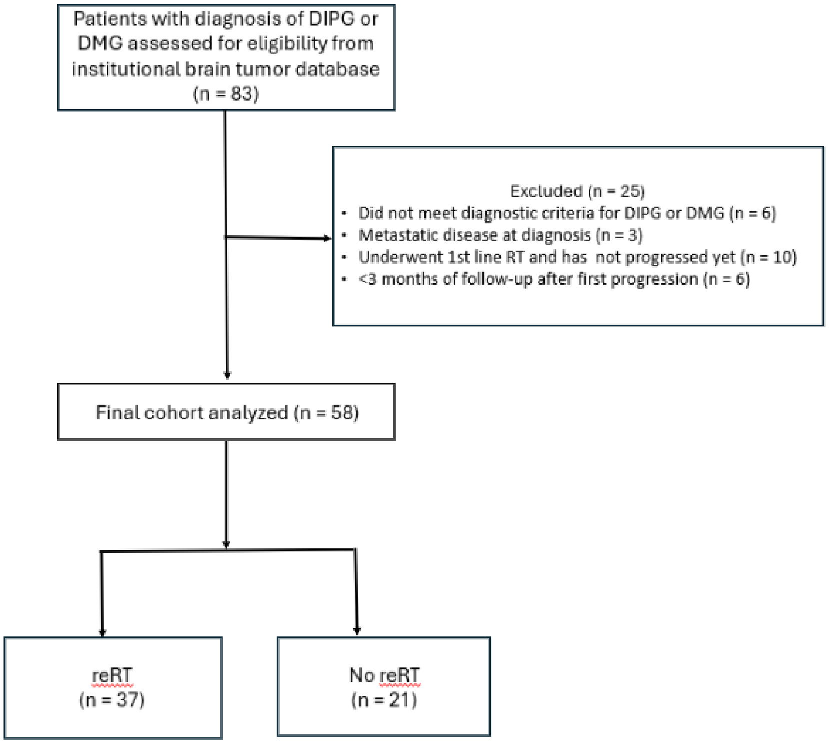


**Supplemental Figure 1.** CONSORT diagram of study screening and cohort selection.

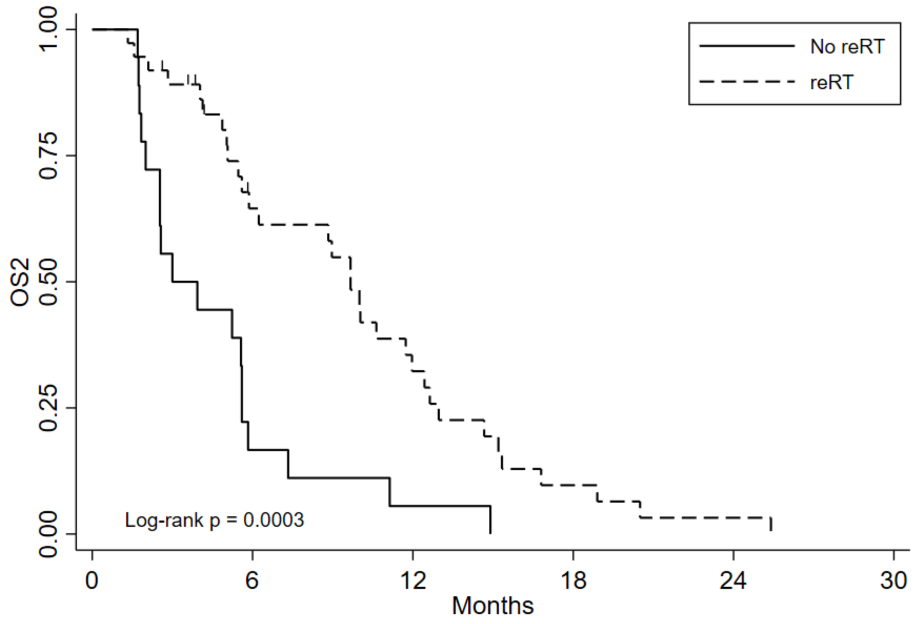


| Number at risk | | | | | | |
|---|---|---|---|---|---|---|
| No reRT | 18 | 3 | 1 | 0 | 0 | 0 |
| reRT | 37 | 20 | 10 | 3 | 1 | 0 |

**Supplemental Figure 2.** Landmark Kaplan–Meier analysis of OS2 stratified by reRT.

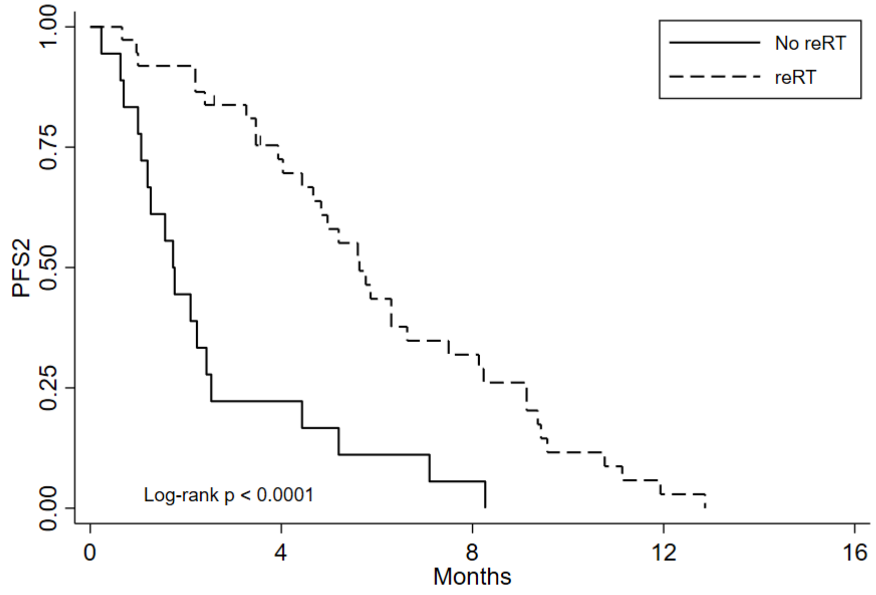


| Number at risk | | | | | |
|---|---|---|---|---|---|
| No reRT | 18 | 4 | 1 | 0 | 0 |
| ReRT | 37 | 25 | 11 | 1 | 0 |

**Supplemental Figure 3.** Landmark Kaplan–Meier analysis of PFS2 stratified by reRT.

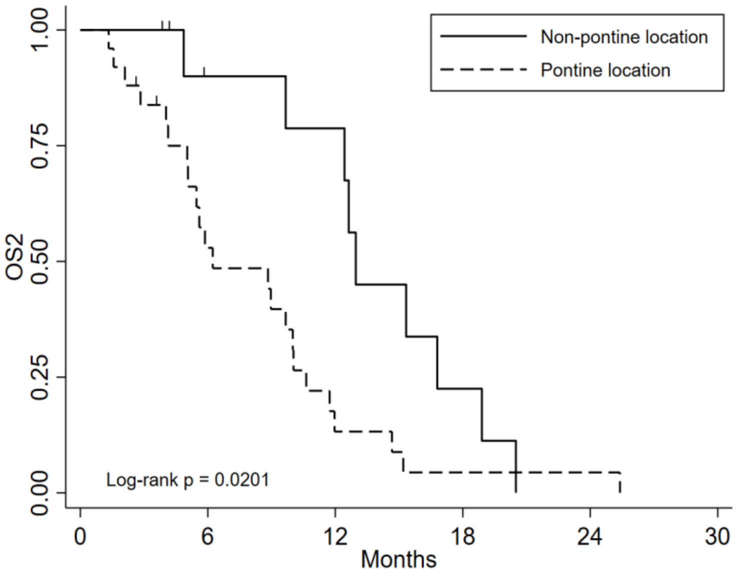


| Number at risk | | | | | | |
|---|---|---|---|---|---|---|
| Non-pontine location | 12 | 8 | 7 | 2 | 0 | 0 |
| Pontine location | 25 | 12 | 3 | 1 | 1 | 0 |

**Supplemental Figure 4.** Kaplan–Meier analysis of OS2 stratified by pontine location among patients who received reRT.

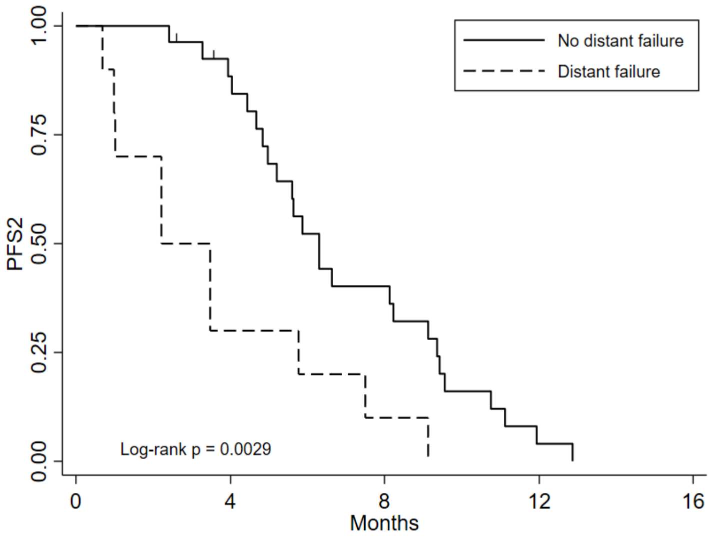


| Number at risk | | | | | |
|---|---|---|---|---|---|
| No distant failure | 27 | 22 | 10 | 1 | 0 |
| Distant failure | 10 | 3 | 1 | 0 | 0 |

**Supplemental Figure 5.** Kaplan–Meier analysis of PFS2 stratified by distant failure among patients who received reRT.
